# Characterizing artificial intelligence (AI) psychosis in a large academic medical setting: evidence of the new clinical phenomenon and the vulnerability of those in early phases of psychosis

**DOI:** 10.64898/2026.06.04.26354939

**Authors:** Zachary Bergson, Sarah G. Vassall, Adam Wright, Allison B. McCoy, Katherine M. Schafer, Margaret C. Achee, Julia M. Sheffield

## Abstract

**Background:** Concerns about “AI psychosis” have swirled in the media since ChatGPT’s release, but few systematic analyses exist. We therefore conducted an electronic health record (EHR) analysis to identify the frequency, clinical characteristics, and quality of AI interactions in patients experiencing psychosis treated in a medical center.

**Methods:** AI keywords (e.g., ChatGPT, AI) were used to search Vanderbilt University Medical Center’s EHR from 12/1/2022-4/1/2026. Records were discarded if they were not AI-related or if the primary diagnosis did not include psychosis. Three raters read notes to determine if a patient was experiencing AI psychosis and classified the interactions using 4 a-priori categories (Catalyst, Amplifier, Co-Author, Object) formulated to explain how AI-related negative outcomes emerge.

**Findings:** 73 patients met our criteria. 28 patients were rated as experiencing AI psychosis, 17 had neutral interactions, and 28 expressed delusional content related to AI without documented evidence of conversational AI use. ChatGPT was the matching keyword for 53.6% patients experiencing AI psychosis. The majority of AI psychosis cases were documented after ChatGPT’s “4o” model was released in May 2024. Notably, the AI Psychosis group had significantly more patients experiencing a first psychotic episode (60.7%) compared to the other two groups. Amplifier was the most common (64.3%) qualitative rating in the AI Psychosis group.

**Interpretation:** “AI psychosis” is an infrequent but real phenomenon observed in clinical practice. Most affected patients were experiencing their first psychotic episode and presented with AI psychosis following the release of the more sycophantic GPT-4o. Among the affected patients, AI most often exacerbated an existing condition by reinforcing distorted ideas.

## Introduction

Conversations about artificial intelligence (AI) and mental health have steadily increased in the media and academic journals since ChatGPT’s public release on November 30, 2022. What started as anecdotal, single case stories^1–5^ reported in the media, has ballooned into an entire (albeit, nascent) field of study. Academic articles on this subject have ranged from viewpoints warning of the dangers of AI providing therapy to individuals with serious mental illness^6,7^ to assessing how AIs detect and manage suicidal ideation in simulated scenarios^8^. Large survey studies have also been conducted and have identified growing use of AI for mental health advice among youths (13.1%), especially among individuals between 18-21 years of age (22.2%)^9^ – a notably vulnerable age for the development of mental illness^10^.

One of the most conspicuous mental health impacts of AI use is psychosis (frequently referred to as “AI Psychosis”), which reflects the development and/or worsening of psychotic symptoms in the context of conversational AI (also called chatbots) use^11^. This has been particularly noted for the development of delusional beliefs (i.e. unfounded beliefs that are not shared by others), with patients, clinicians and researchers noting how chatbots can serve as reinforcers of unusual thought content^12–15^. There is growing concern that engagement with chatbots, which leverage large language models trained on vast amount of text to address user prompts^16^, may expand upon or even create delusion beliefs in the mind of some users, leading to documented consequences of hospitalization, social dysfunction, and arrest^13^.

However, to date, research on AI Psychosis has been limited to case studies, theoretical viewpoints, and simulated experimental studies^6,7,11,14,15,17,18^. While critical and helpful starting points, there is a lack of empirical data regarding the characteristics of individuals who develop AI Psychosis, the frequency of their presentation to clinical settings, and aggregate data on the impact of AI on their mental health. For instance, many high-profile case stories described by the media have focused on individuals with no clear psychiatric history^19–22^ while clinicians have anecdotally noted increasing cases of patients engaging with these products in ways that worsen pre-existing psychopathology^12^. Therefore, it is vital to elucidate the clinical and demographic factors characteristic of those who present with AI Psychosis, to begin understanding the true nature of vulnerability to its development. Additionally, mental health clinicians would greatly benefit from a systematic analysis to determine if these interactions are appearing in common care environments (e.g., hospitals, outpatient clinics) and, if so, how they are manifesting in patients.

To this end, we conducted a retrospective electronic health record (EHR) analysis of documented cases of AI interactions at a large medical center. Our aims were threefold: first, to estimate prevalence of AI Psychosis amongst those presenting for clinical care; second, to identify clinical and demographic features of affected patients; third, to characterize the impact of conversational AI on psychotic symptoms.

## Methods

We conducted this study at Vanderbilt University Medical Center (VUMC), a large medical center in Nashville, Tennessee. It was approved by the VUMC Institutional Review Board (IRB #251906) with a waiver of informed consent, as it involved retrospective analysis of data collected during routine clinical care and posed no more than minimal risk to participants.

### Data collection and screening

History of presenting illness (description of patient’s present condition) and progress notes were extracted from the EHR using Epic’s Clarity tool^23^. Records from December 1, 2022 to April 1, 2026 were included if they contained the following keywords: AI, Chatbot, ChatGPT, Claude, Gemini, Grok, Copilot, Perplexity, Llama, or Bard. Only patients who received services (before or after December 1, 2022) from a mental health provider (psychiatrist, psychologist, social worker, medical resident, etc.) at VUMC were queried in this keyword search. Patient demographic information of age, sex, date of encounter, and race were recorded for each identified record. Given our focus on AI Psychosis, any psychosis-related diagnosis was also recorded in the first Epic Clarity search: schizophrenia, schizoaffective disorder, schizophreniform disorder, brief psychotic disorder, delusional disorder, bipolar disorder with psychotic features, major depressive disorder with psychotic features, unspecified psychosis, and substance-induced psychosis. This allowed us to capture patients who had clinically-meaningful psychosis, as opposed to those with unusual or eccentric beliefs that were not impacting their mental health.

After these records were collected, the data were additionally screened using a keyword search for the following psychiatric terms: psychiatry, psychology, schizophrenia, psychosis, grandiose, grandiosity, delusion, hallucination, paranoia, bipolar, and mania. Records that did not include one of these terms were excluded from the analysis. Patients without a psychotic disorder in their chart were excluded.

The note text of the remaining records was then manually screened by authors ZB and SGV. Records were discarded if the AI keyword match was incidental (e.g., patient name) or if the record lacked a relevant psychotic disorder diagnosis upon further record review. The diagnostic code assigned to the encounter was used as the patient’s primary diagnosis. When more than one record was present for a patient, the most recent note’s diagnostic label was used.

Presence or absence of different aspects of the patient’s psychiatric history preceding the documented AI interaction were then manually coded based on a review of each patient’s EHR: prior psychiatric hospitalization, history of prescribed psychotropic medication, history of psychiatric outpatient treatment (psychotherapy, medication management), and history of suicide attempts. These characteristics were readily available in the EHR, as they are included in all psychiatric history of presenting illness (H&P) notes and initial assessments. Furthermore, chart review was conducted (ZGB, JMS, and MA) to determine if the documented AI interaction was part of their first episode of psychosis (“index episode”). This was defined as the AI interaction being documented by their treatment team during their first presentation to psychiatric services for psychotic symptoms (e.g. if the patient presented to the hospital for treatment of their first episode psychosis, and the AI interaction was documented on day 4 of that hospitalization, it would be considered part of their index episode). First episode was defined as the *first* seeking of treatment for psychosis in the patient’s lifetime.

Finally, text from the relevant EHR notes was used by authors ZB and SGV to categorize LLM interactions according to the following schema: 1) neutral AI interaction (e.g., using ChatGPT to search for recipes, did not negatively interact with the psychosis); 2) AI Psychosis (e.g., AI use appeared to negatively interact with or exacerbate the psychosis; or 3) AI-related psychotic content (e.g., AI as central to the delusion without clear evidence of the patient using conversational AI). The final category was included to ensure that individuals were captured who know of AI and incorporate it into their delusion framework (e.g., “AI has replaced my family”), but who may not have directly interacted with these products and/or had their psychosis worsened by the *interaction* with these products. In the case of rater disagreement, a tiebreaking rater was used for the final determination (JMS).

### AI Psychosis category screening

Recent theoretical work has described four *a priori* categories to typify different AI Psychosis presentations^13^. Authors ZB, MA, and JMS, clinical psychologists with extensive experience treating and assessing individuals with psychotic disorders, familiarized themselves with the AI Psychosis categories (Catalyst, Amplifier, Co-Author, and Object) and assigned their best estimate category for the interaction described in the note. Figure 1 provides brief descriptions of these four categories. An additional category, Other, was added as an option if the raters felt a case did not fit into the four *a priori* options. ZB and MA examined note text and reviewed each patient’s EHR (e.g., examining psychiatric and social history, information from family, follow up interventions) to assign a rating. JMS served as a third rater to tiebreak disagreements. If a patient had more than one record, their most common rating was used in the descriptive analysis.

**Figure 1.**
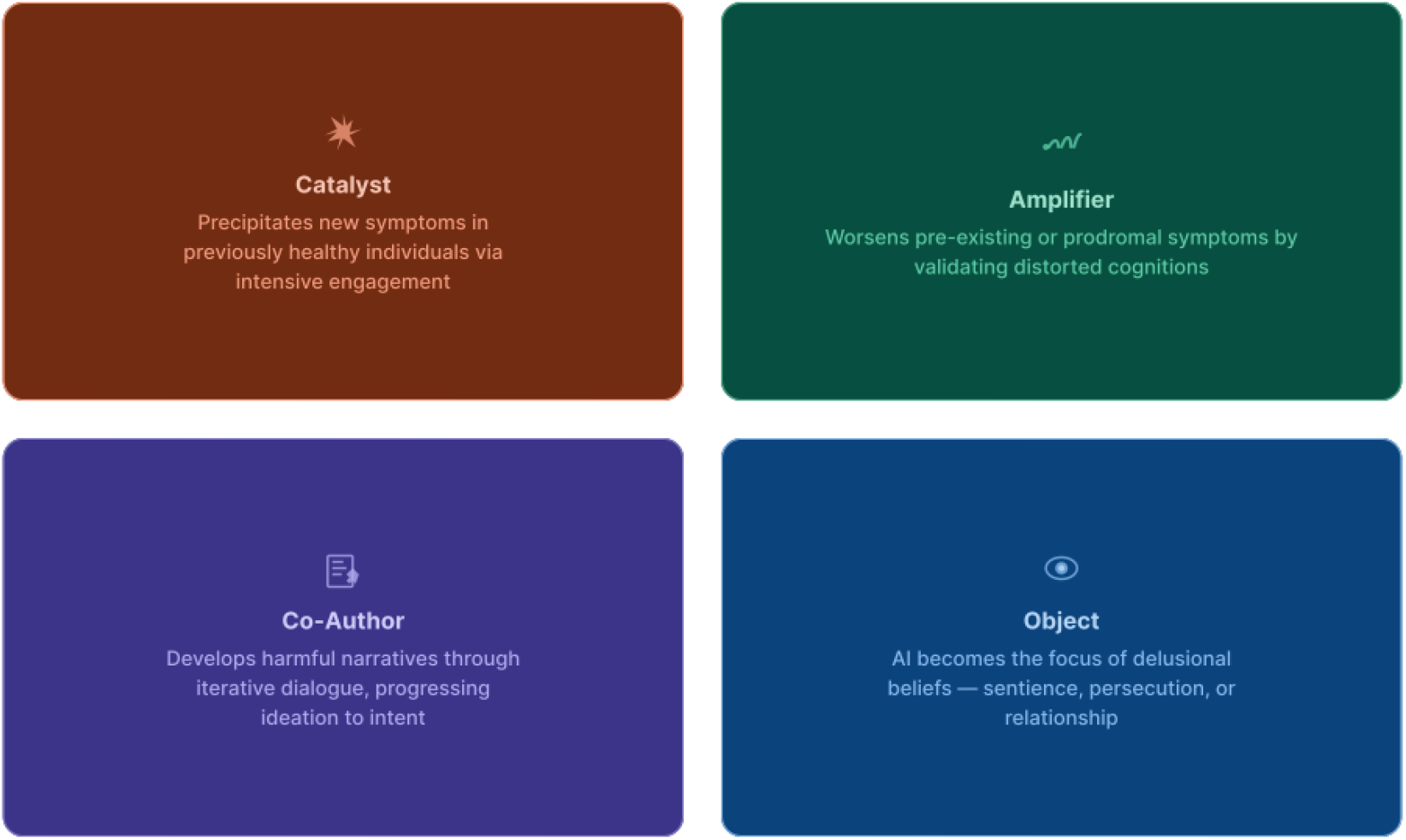
Descriptions of AI Psychosis categories created by Flathers, Roux, and Torous (2026).

### Descriptive and statistical analyses

Jamovi^18^ and R^17^ were used to complete all data visualizations and analyses. Participants with a diagnosis of a psychotic disorder were rated by type of AI interaction (neutral, AI Psychosis, AI-related psychotic content); inter-rater reliability was calculated as percent agreement between raters across all eligible encounters. Across the three groups, we compared age using a Kruskal-Wallis test; chi-square tests of independence were used to compare the three groups on sex, race, primary diagnosis, and relevant psychiatric history (i.e., history of prior hospitalizations, psychotropic medication use, outpatient therapy, and suicide attempts). For significant chi-square tests, pairwise comparisons were conducted, with holm correction. Of those who had AI Psychosis, the role of conversational AI in psychosis worsening was rated by clinicians (Catalyst, Amplifier, Co-Author, and Object) and inter-rater reliability was again calculated as percent agreement between raters.

## Results

### Electronic health record search

From December 1, 2022 to April 1, 2026, a total of 578,058 records (215,712 unique patients) occurred across all VUMC psychiatry and psychology services and departments; of these, 21,665 records met initial search term inclusion criteria (Figure 2). However, the keywords “Claude”, “Perplexity”, and “Copilot” appeared frequently but were discarded during manual screening: “Claude” overlapped exclusively with patient names; “Perplexity” matched a common psychiatric descriptor; and “Copilot” overlapped with an EHR documentation tool. The keyword “AI” also often matched providers discussing autoimmune conditions and was frequently— although not always—discarded.

**Figure 2.**
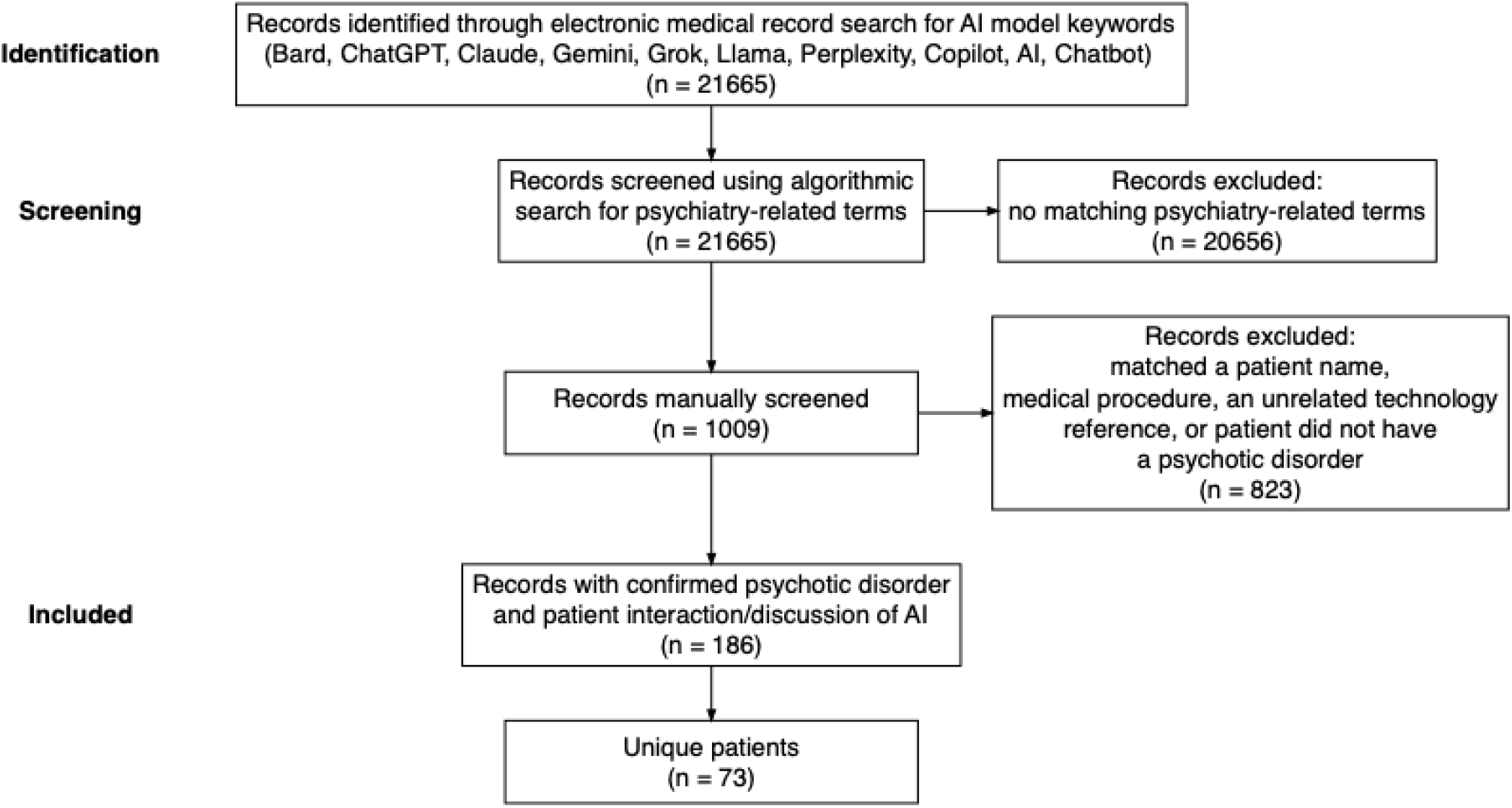
Electronic health record screening workflow

After algorithmic and manual screening was completed, 186 encounters across 73 unique patients remained. “AI” was the most prominent keyword match, capturing 141 encounters (81.5%), followed by “ChatGPT”, which matched 44 encounters (25.4%). “Gemini” had 1 matching encounter (0.57%). 53.6% of AI Psychosis patients were documented as using ChatGPT, 3.6% were using Gemini, and the rest (42.8%) were described as using a conversational AI product that was not specifically documented by the provider. Psychiatrists, psychologists, social workers, licensed professional counselors, and neurologists comprised the providers for the encounters included in our final dataset.

Percent agreement for whether the AI interaction was considered neutral, psychosis-worsening, or AI-related psychotic content was 86%.

### Demographics and clinical characteristics of the AI Psychosis cohort

Among this 73-patient sample, 28 were labeled as having psychosis-worsening interactions with AI, henceforth termed “AI Psychosis” (Table 1). These 28 cases represented 0.013% of individuals seen for mental health care at VUMC during this period.

**Table 1.**
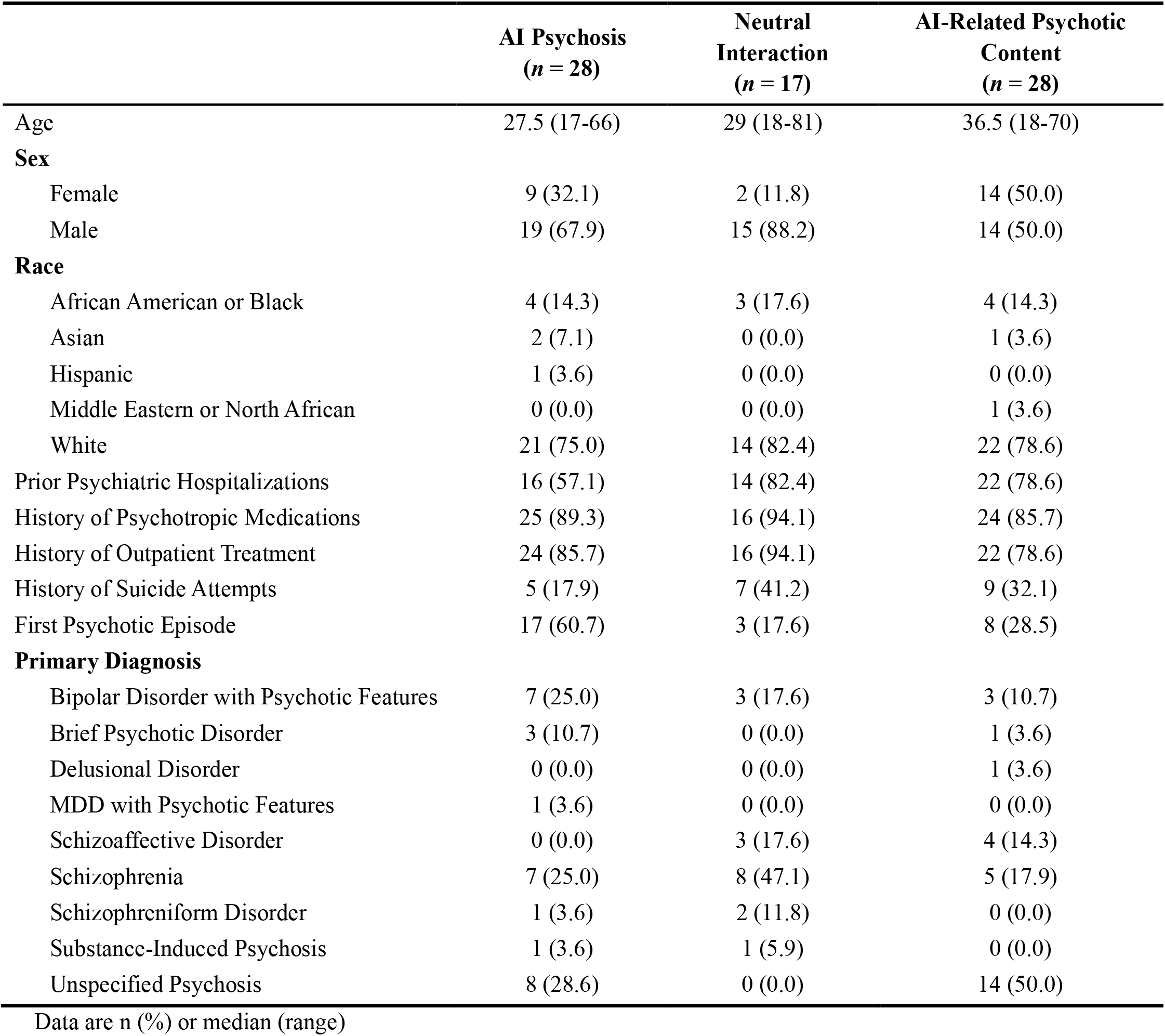
Demographic and clinical characteristics.

|  | **AI Psychosis**<br>**(n = 28)** | **Neutral**<br>**Interaction**<br>**(n = 17)** | **AI-Related Psychotic**<br>**Content**<br>**(n = 28)** |
| --- | --- | --- | --- |
| Age | 27.5 (17-66) | 29 (18-81) | 36.5 (18-70) |
| **Sex** |  |  |  |
| Female | 9 (32.1) | 2 (11.8) | 14 (50.0) |
| Male | 19 (67.9) | 15 (88.2) | 14 (50.0) |
| **Race** |  |  |  |
| African American or Black | 4 (14.3) | 3 (17.6) | 4 (14.3) |
| Asian | 2 (7.1) | 0 (0.0) | 1 (3.6) |
| Hispanic | 1 (3.6) | 0 (0.0) | 0 (0.0) |
| Middle Eastern or North African | 0 (0.0) | 0 (0.0) | 1 (3.6) |
| White | 21 (75.0) | 14 (82.4) | 22 (78.6) |
| Prior Psychiatric Hospitalizations | 16 (57.1) | 14 (82.4) | 22 (78.6) |
| History of Psychotropic Medications | 25 (89.3) | 16 (94.1) | 24 (85.7) |
| History of Outpatient Treatment | 24 (85.7) | 16 (94.1) | 22 (78.6) |
| History of Suicide Attempts | 5 (17.9) | 7 (41.2) | 9 (32.1) |
| First Psychotic Episode | 17 (60.7) | 3 (17.6) | 8 (28.5) |
| **Primary Diagnosis** |  |  |  |
| Bipolar Disorder with Psychotic Features | 7 (25.0) | 3 (17.6) | 3 (10.7) |
| Brief Psychotic Disorder | 3 (10.7) | 0 (0.0) | 1 (3.6) |
| Delusional Disorder | 0 (0.0) | 0 (0.0) | 1 (3.6) |
| MDD with Psychotic Features | 1 (3.6) | 0 (0.0) | 0 (0.0) |
| Schizoaffective Disorder | 0 (0.0) | 3 (17.6) | 4 (14.3) |
| Schizophrenia | 7 (25.0) | 8 (47.1) | 5 (17.9) |
| Schizophreniform Disorder | 1 (3.6) | 2 (11.8) | 0 (0.0) |
| Substance-Induced Psychosis | 1 (3.6) | 1 (5.9) | 0 (0.0) |
| Unspecified Psychosis | 8 (28.6) | 0 (0.0) | 14 (50.0) |

Data are n (%) or median (range)

Regarding patient characteristics, the AI Psychosis cohort was majority male (67.9%) and majority white (75%). Unspecified psychosis was the most common diagnostic label given to AI Psychosis patients (28.6%), followed by bipolar disorder with psychotic features (25%) and schizophrenia (25%). The majority of the AI Psychosis group (60.7%) were experiencing their first psychotic episode.

Of those with AI Psychosis, 57.1% had experienced at least one psychiatric hospitalization, 32.1% had been hospitalized prior to the AI Psychosis episode, 17.9% had a history of suicide attempt, and 25 of 28 patients (89.2%) had been prescribed psychotropic medications before their documented AI Psychosis interaction (Figure 3). 24 of the 28 (86%) AI Psychosis interactions were documented after the release of ChatGPT’s GPT-4o model in May 2024 (Figure 4), which had new features that led to more sycophantic interactions with users^24^. The majority of patients with AI-related psychotic content or neutral interactions were also documented by providers after GPT-4o was released (Supplementary Figures 1-2).

**Figure 3.**
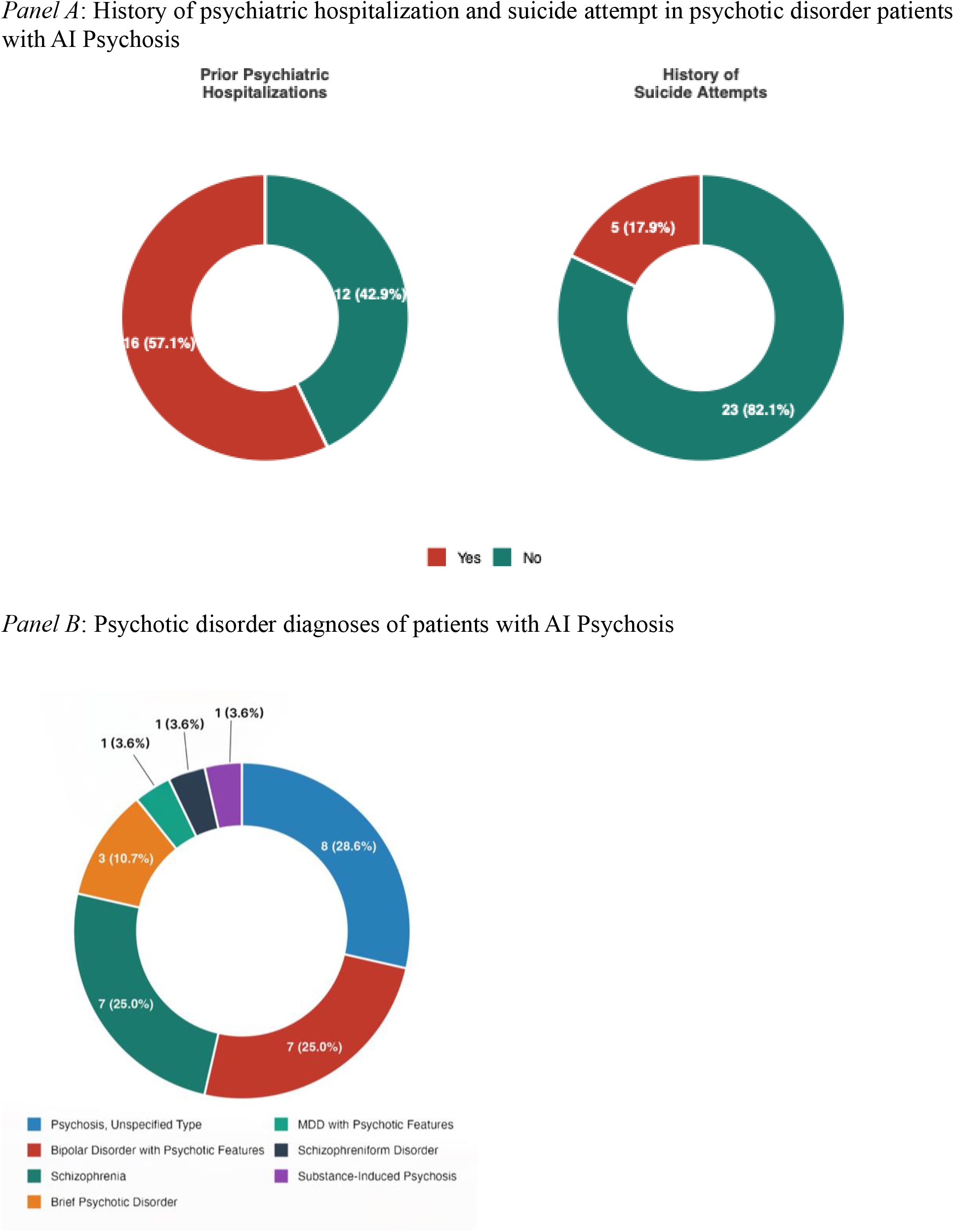
Clinical characteristics of psychotic disorder patients with psychosis-worsening AI interactions (*n* = 28)

**Figure 4.**
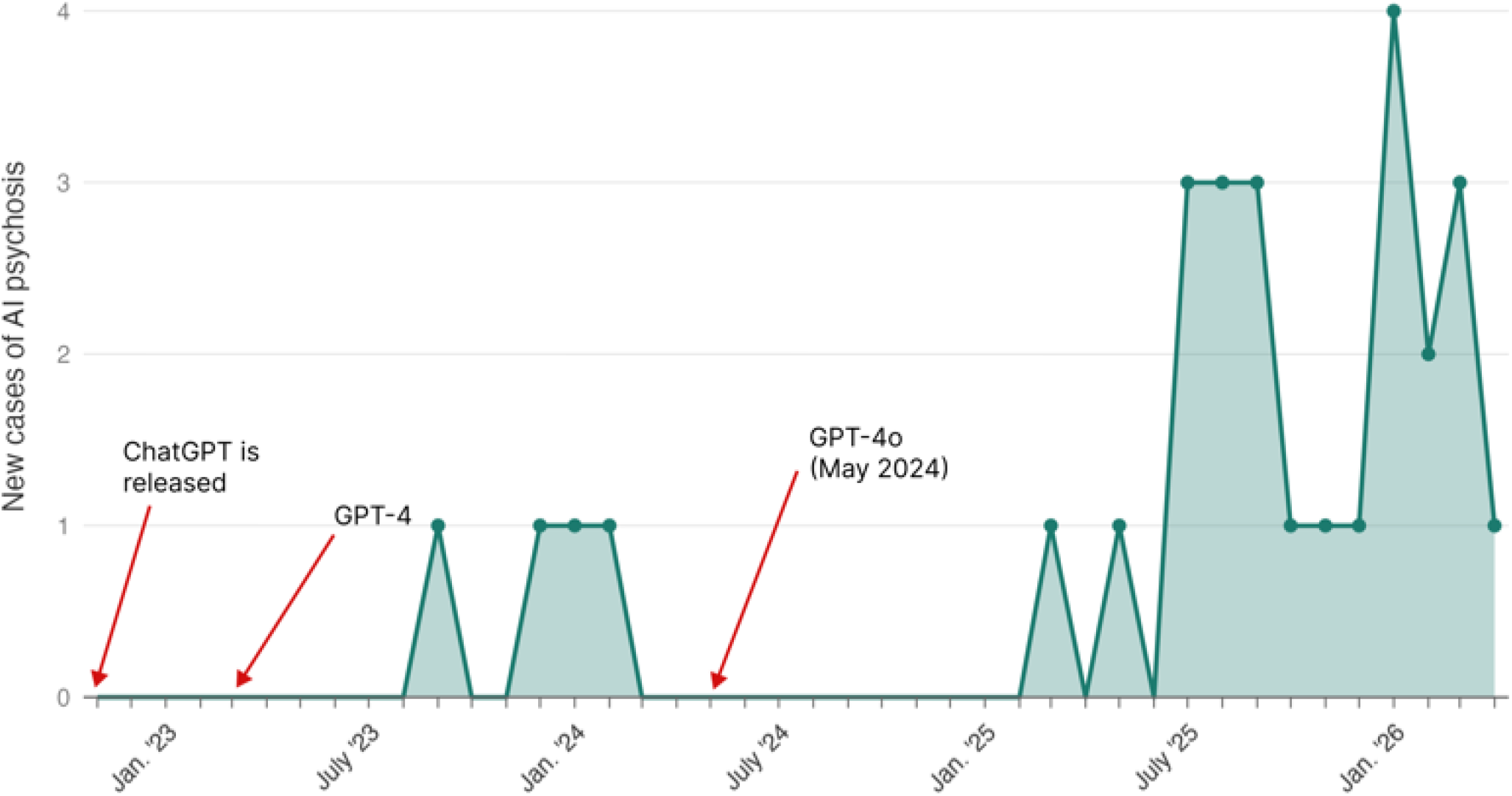
Tracking the occurrence of AI Psychosis in patients following ChatGPT’s release on November 30, 2022 (*n* = 28)

### Comparison with other groups

AI Psychosis patients did not significantly differ from those with neutral interactions or AI-related psychotic content on any clinical history variables such as presence/absence of psychiatric hospitalization, psychotropic use, or suicide attempts (Supplementary Table 1). AI Psychosis patients were significantly younger than the AI-related psychotic content group and were significantly more likely to have an unspecified psychosis diagnosis compared to patients in the neutral group (Supplementary Table 2). There were no significant differences between groups for race or sex after adjusting for multiple comparisons.

Critically, AI Psychosis patients were significantly more likely to be experiencing their first psychotic episode (60.7%) compared to the neutral interaction group (17.6%), or the AI-related psychotic content group (28.5%) (p<.05) (Figure 5; Supplementary Tables 1-2).

**Figure 5.**
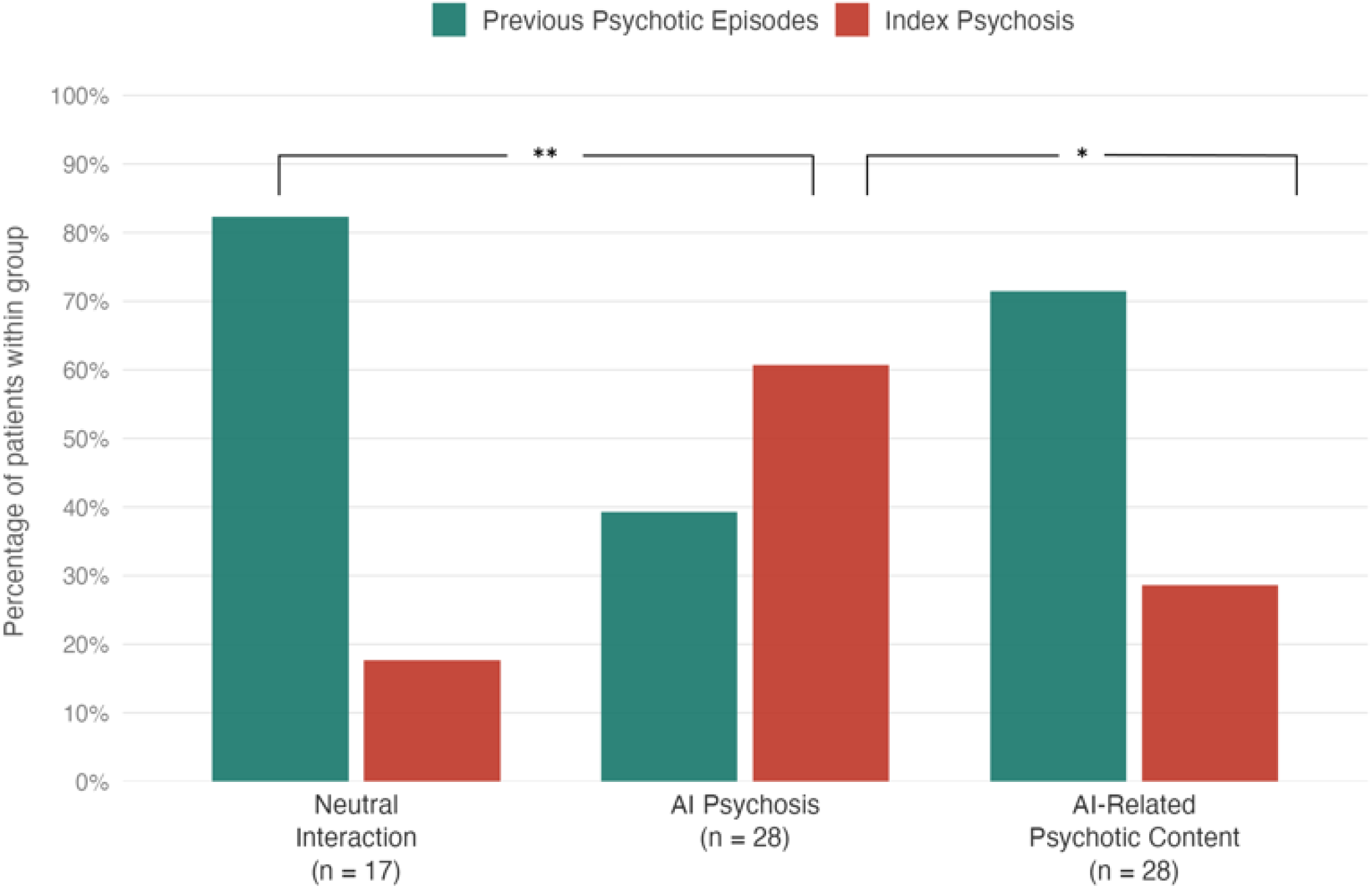
Percentage of patients experiencing first episode (index) psychosis alongside a documented interaction with AI

### Qualitative ratings of AI Psychosis category

Percent agreement for AI Psychosis category was 83.1%. Amplifier was the most common rating assigned (64.3%), followed by Object (21.4%) and Catalyst (10.7%) (Figure 6). Co-Author was not assigned to any record.

**Figure 6.**
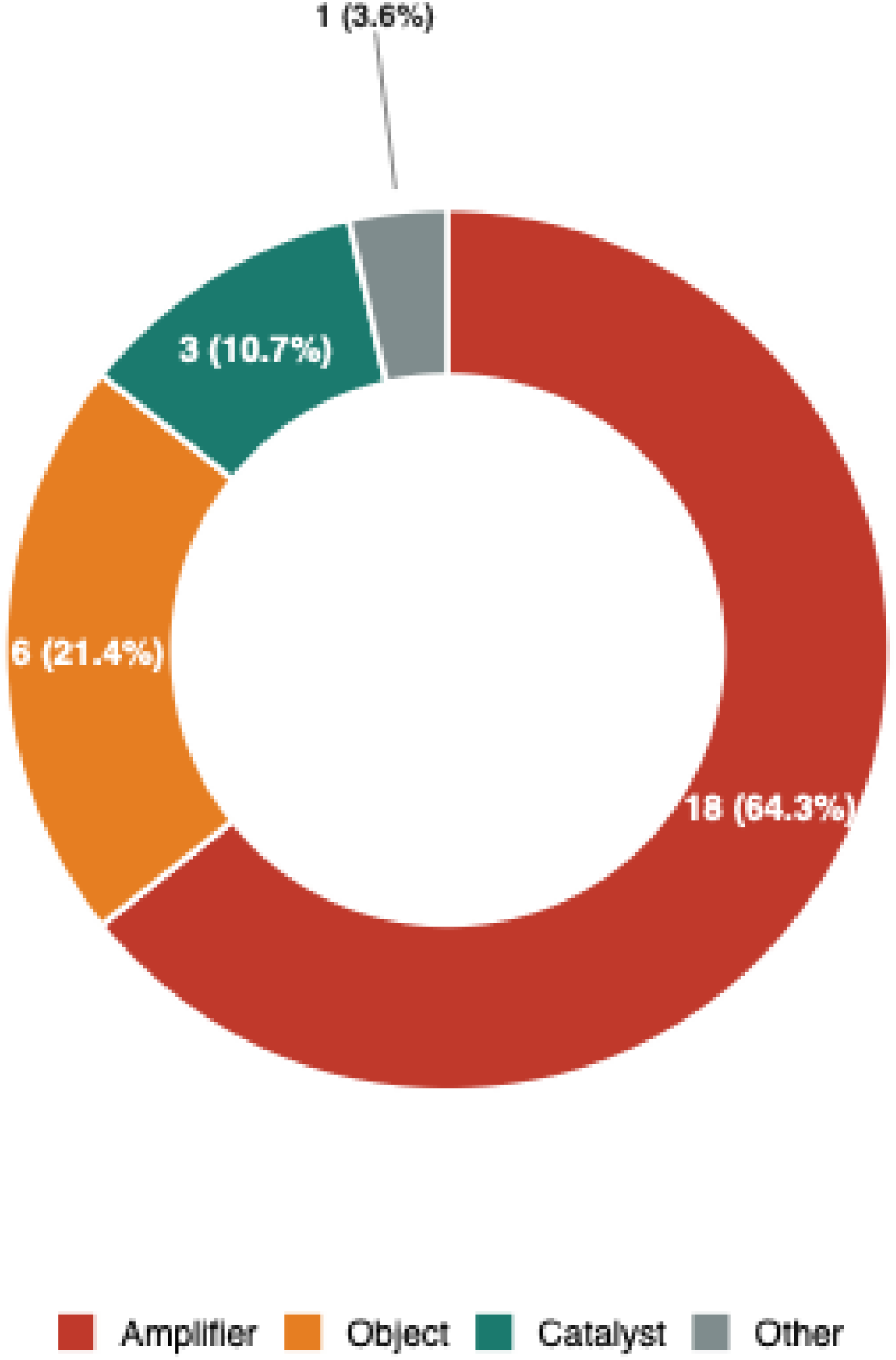
Qualitatively rated AI Psychosis categories in psychotic disorder patients represented in the electronic health record (*n* = 28)

Among individuals with the Amplifier rating, 33.3% had schizophrenia, 27.78% bipolar disorder with psychotic features, 22.2% unspecified psychosis, 5.5% brief psychotic disorder, and 5.5% substance-induced psychosis. In the Object category, 50% had unspecified psychosis, 33.3% bipolar with psychotic features, and 16.7% brief psychotic disorder. Finally, the catalyst category (*n* = 3) was made up of individuals with brief psychotic disorder, unspecified psychosis, or major depressive disorder with psychotic features.

## Discussion

In our retrospective analysis of electronic health records in a large medical center in the United States, we identified 28 documented cases of AI Psychosis since the first major commercial AI chatbot’s (ChatGPT) release in late 2022. This dataset – representing the first aggregate data of documented AI Psychosis in clinical settings -- provides additional context for recent case reports and vignettes by clearly establishing that patients are describing (and/or demonstrating) to providers that the use of AI chatbots is exacerbating psychotic symptoms, confirming AI Psychosis as a genuine clinical phenomenon. Of the affected patients, 53.6% endorsed using ChatGPT, with the majority of all cases occurring after the release of ChatGPT’s popular “4o” model. Many of the AI Psychosis cases were not naïve to psychiatric care; most had been previously prescribed psychiatric medication, and 57.1% (*n*=16) had been previously psychiatrically hospitalized. However, we also observed a striking pattern: compared with patients who had neutral interactions with AI chatbots or incorporate AI technology into their delusions, those with AI Psychosis were significantly younger and were significantly more likely to be experiencing their first psychotic episode. Within this AI Psychosis cohort, Amplifier was the most common type of AI Psychosis interaction, suggesting that AI tools most often intensified emerging or pre-existing psychotic experiences and/or unusual thought content.

Our finding of AI Psychosis cases at a large medical center converges with accumulating evidence showing the risk that conversational AIs pose to mental health^1,12–14,25^. OpenAI, the proprietor of ChatGPT, has publicly acknowledged that “0.07% of users active in a given week and 0.01% of messages indicate possible signs of mental health emergencies related to psychosis or mania”^25^. This converges with our data, which revealed that approximately 0.01% of patients with a history of receiving psychiatric care at VUMC met our criteria for AI Psychosis in clinical encounters from December 1, 2022 to April 1, 2026. Although these rates are small, in a weekly user base of 900 million and growing^26^, the number of negatively affected ChatGPT users (∼630,000) is not insignificant. ChatGPT is reportedly used by more than 75% of monthly conversational AI users^27^.

Our 28 AI Psychosis cases likely underestimate the true frequency. In order to meet inclusion criteria for AI Psychosis in our study, patients needed to have a documented psychotic disorder diagnosis, documented use of AI by providers, and a description in the chart that was convincing of AI use specifically worsening a psychotic experience. This more conservative approach was used to minimize false positives, but these criteria, in addition to our dependence on patients disclosing AI use and providers documenting it, likely missed true cases. For instance, the 28 patients with AI-related psychotic content were classified on the basis of delusions or hallucinations *about* AI without documented use. It is plausible, however, that some of these patients were in fact using AI products in ways that were never disclosed or charted.

In a 2025 blog post reporting mental health-related interactions among weekly users^25^, OpenAI claimed that improvements made to its GPT-5 model, the successor to GPT-4o, reduced “undesired responses” to severe mental health symptoms like mania and psychosis by 39% as judged by consulting psychiatrists and psychologists. This change was made after findings that GPT-4o was sycophantic, unlikely to challenge users, and responded inappropriately to psychotic prompts^7,18,24,28^, a notably harmful and exacerbating approach for reinforcing delusional beliefs^29^. The incidence of new AI Psychosis cases at VUMC, however, does not indicate a reduction in negative interactions since GPT-5’s release on August 7, 2025 (Figure 4). This could be explained by the growing popularity of ChatGPT naturally increasing the incidence rate of AI Psychosis, growing clinician awareness of AI Psychosis, OpenAI’s delay in removing GPT-4o from the market until January 29, 2026^30^, or affected patients (42.8% for whom a specific AI product wasn’t documented) using other conversational AI products with poor mental health safety. It is also possible, however, that sycophancy and other safety concerns have not been fully trained out of ChatGPT.

Clinically, AI Psychosis patients were similar to the other two groups in our sample with regards to broad aspects of their psychiatric experiences (e.g., hospitalization; psychotropic medication). History of suicide attempt in the AI Psychosis group was 17.9%, which is higher than prevalence rates for individuals with schizophrenia (10%)^31^ and lower than those with bipolar disorder (30-60%)^32^. Notably, 60.7% were experiencing their first psychotic episode, significantly more than the other two groups, suggesting that AI Psychosis may occur more frequently in the early phases of psychotic illness. This is further supported by the significantly younger age of these patients and the fact that 28.6% of affected patients were diagnosed with “unspecified psychosis” and 10.3% with brief psychotic disorder. Unspecified psychosis is often assigned to individuals in the early stages of psychotic illness when psychiatric history preceding the psychotic episode is unclear^33^. Brief psychotic episodes are associated with high risk for developing a more chronic psychotic disorder^34^. These findings suggest chatbot use may confer more risk to patients in the early stage of psychotic illness.

Finally, the preponderance of Amplifier cases (64.3%) in our study provides the field with preliminary evidence that AI Psychosis largely manifests as a worsening or exacerbation of emerging or pre-existing psychotic symptoms. This pattern is concerning given predictive processing accounts of delusions, which state that new information (e.g. from a chatbot conversation) is used to strengthen delusional priors, further biasing information processing towards confirming the delusional beliefs^35,36^. Although we lacked access to patients’ chatlogs, Amplifier patients may have been discussing unusual beliefs with conversational AI, asking for help making sense of odd thoughts or experiences (e.g. noticing coincidences or patterns in their environment), that are typical of the early stages of psychosis exacerbation^37,38^. Within this framework, AI validation and elaboration of such beliefs could plausibly entrench sticky priors of a volatile or dangerous world, making them more resistant to updating with disconfirming evidence^35^.

More research using prospective study designs is clearly needed to estimate true prevalence and to understand the occurrence, clinical implications, and phenomenology of AI Psychosis. Without direct access to chat logs in tandem with detailed clinical histories, it will be difficult for psychiatric researchers to fully understand how AI Psychosis emerges. Is excessive use of these platforms contributing to sleep dysfunction, a significant risk factor for psychotic disorders^39^? Are the AIs failing to challenge distorted thoughts^13,40^? Or is it taking individuals down unhelpful lines of thought and inquiry in order to keep them engaged? A dearth of evidence exists at this time, and may require researchers to either gain access to chat logs with affected individuals’ consent and/or develop direct partnerships with companies like OpenAI, Anthropic, and Google.

There are several limitations to our retrospective EHR analysis. Ascertainment bias likely affected case identification: detection required patient disclosure, clinician recognition, quality of the documentation, and keyword capture, with failure at any step potentially yielding a missed or misclassified case. Our keywords were also necessarily incomplete, as the list of available consumer AI products has grown extensively since ChatGPT’s release. Findings are further limited by the single-site design and use of a theoretically-informed, but not clinically-validated, AI Psychosis rating system (i.e., the Co-Author rating was not assigned to any cases).

In conclusion, this retrospective analysis offers initial evidence that AI Psychosis is a true clinical phenomenon reported by patients and documented by psychiatric providers. Patients in the early phases of psychotic illness may be particularly vulnerable to interactions with conversational AI that exacerbate unusual thoughts and experiences. Amplification of emerging or pre-existing symptoms appears to be the predominant type of AI Psychosis, with a smaller subset of cases consistent with de novo psychosis, in which AI use was considered the catalyst for the psychotic symptoms. Routine assessment of AI use in psychiatric encounters, alongside prospective research into mechanisms and clinical correlates, is critical as conversational AI continues to expand.

## Data Availability

Data for this study cannot be shared due to HIPPA concerns involving the distribution of personal health information.

## Declaration of interests

The authors do not have conflicts of interest related to this study.

## Acknowledgements

This work was supported by the Charlotte and Donald Test Fund, the Vanderbilt Institute for Clinical and Translational Research (Grant No. VR74021), and the National Institute of Mental Health (Grant No. 1R01MH137024-01)

## Notes

### Competing Interest Statement

The authors have declared no competing interest.

### Author Declarations

The IRB of Vanderbilt University Medical Center gave ethical approval for this work (IRB #251906).

