## Supplemental Tables and Figures for "Characterizing artificial intelligence (AI) psychosis in a large academic medical setting: evidence of the new clinical phenomenon and the vulnerability of those in early phases of psychosis"

**Supplementary Table 1.** Demographic and clinical statistical comparisons between patients with AI psychosis neutral AI interactions, and AI-related psychotic content

| | **AI Psychosis (n = 28)** | **Neutral Interaction (n = 17)** | **AI-related psychotic content (n = 28)** | $\chi^2$ **(df)** | ***p*** |
|---|---|---|---|---|---|
| Age* | 27.5 | 29 | 36.5 | 9.29 (2) | .009 |
| **Sex** | | | | 6.95 (2) | .031 |
| Female | 9 (-0.29) | 2 (-2.23) | 14 (2.23) | | |
| Male | 19 (0.29) | 15 (2.23) | 14 (-2.23) | | |
| **Race** | | | | 4.73 (8) | .786 |
| African American | 4 | 3 | 4 | | |
| Asian | 2 | 0 | 1 | | |
| Hispanic | 1 | 0 | 0 | | |
| Middle Eastern or North African | 0 | 0 | 1 | | |
| White | 21 | 14 | 22 | | |
| Prior Psychiatric Hospitalizations | 16 | 14 | 22 | 4.47 (2) | .107 |
| History of Psychotropic Medications | 25 | 16 | 24 | 0.77 (2) | .681 |
| History of Outpatient Treatment | 24 | 16 | 22 | 2.02 (2) | .364 |
| History of Suicide Attempts | 5 | 7 | 9 | 3.06 (2) | .217 |
| First Psychotic Episode | 11 (-3.09) | 14 (2.0) | 20 (1.36) | 10.1 (2) | .006 |
| **Primary Diagnosis** | | | | 29.1 (16) | .024 |
| BD with Psychotic Features | 7 (1.26) | 3 (-0.02) | 3 (-1.25) | | |
| Brief Psychotic Disorder | 3 (1.55) | 0 (-1.33) | 1 (-0.56) | | |
| Delusional Disorder | 0 (-0.79) | 0 (-0.55) | 1 (1.28) | | |
| MDD with Psychotic Features | 1 (1.28) | 0 (-0.55) | 0 (-0.79) | | |
| Schizoaffective disorder | 0 (-2.19) | 3 (1.29) | 4 (1.08) | | |
| Schizophrenia | 7 (-0.36) | 8 (2.07) | 5 (-1.44) | | |
| Schizophreniform Disorder | 1 (-0.18) | 2 (1.82) | 0 (-1.39) | | |
| Substance Induced Psychosis | 1 (0.34) | 1 (0.91) | 0 (-1.13) | | |
| Unspecified Psychosis | 8 (-0.22) | 0 (-3.09) | 14 (2.92) | | |

Data are n or median, and data in parentheses are chi-square ($z_{ij}$) standardized residuals. Standardized residuals were not calculated for analyses with a $p > .05$ for its omnibus test. All statistical tests are chi-square analyses unless otherwise specified.

* A Kruskal-Wallis one-way ANOVA (dependent variable, age, did not meet normality assumptions) was run to examine age differences among the 3 groups $\chi^2(2) = 9.39$, $p = .009$. Post hoc comparisons (Dwass-Steel-Critchlow) revealed the AI psychosis group was significantly younger than the AI-related psychotic content group $W$=3.931, $p = .015$. No significant differences were identified in the other pairwise comparisons.

**Table 2.** Post-hoc pairwise comparisons of sex, psychosis history, and primary diagnosis across AI interaction rating groups

| | Pairwise rating-group comparisons ($p$, $p_{adj}$) | | |
|---|---|---|---|
| | **Neutral vs AI Psychosis** | **Neutral vs AI-related psychotic content** | **AI Psychosis vs AI-related psychotic content** |
| **Sex** | | | |
| Female | .236, .472 | .023, .068 | .277, .472 |
| Male | .236, .472 | .023, .068 | .277, .472 |
| First Psychotic Episode | .012, .036 | .639, .639 | .031, .063 |
| **Diagnosis** | | | |
| Schizophrenia | .232, .464 | .079, .237 | .745, .745 |
| Schizoaffective Disorder | .092, .276 | .999, .999 | .12, .276 |
| Bipolar Disorder with Psychotic Features | .837, .999 | .833, .999 | .295, .886 |
| Psychosis, Unspecified Type | .043, .085 | .001, .004 | .171, .171 |
| Brief Psychotic Disorder | .435, .999 | .999, .999 | .604, .999 |
| Schizophreniform Disorder | .651, .999 | .267, .8 | .999, .999 |
| Delusional Disorder* | — | — | — |
| MDD with Psychotic Features* | — | — | — |
| Substance-Induced Psychosis* | — | — | — |

*Note.* Pairwise comparisons of category proportions (each category vs. all others) using Fisher's exact test. $p$ adjusted using Holm's method. Omnibus tests and standardized residuals are reported in Table 1.

* Pairwise comparisons could not be calculated for these diagnoses due to small size ($n$ = 1 or $n$ = 2)

**Supplementary Figure 1.** Tracking the documentation of AI-related psychotic content (with no clear interaction with an AI model) in patients following ChatGPT's release on November 30, 2022 ($n$ = 28)

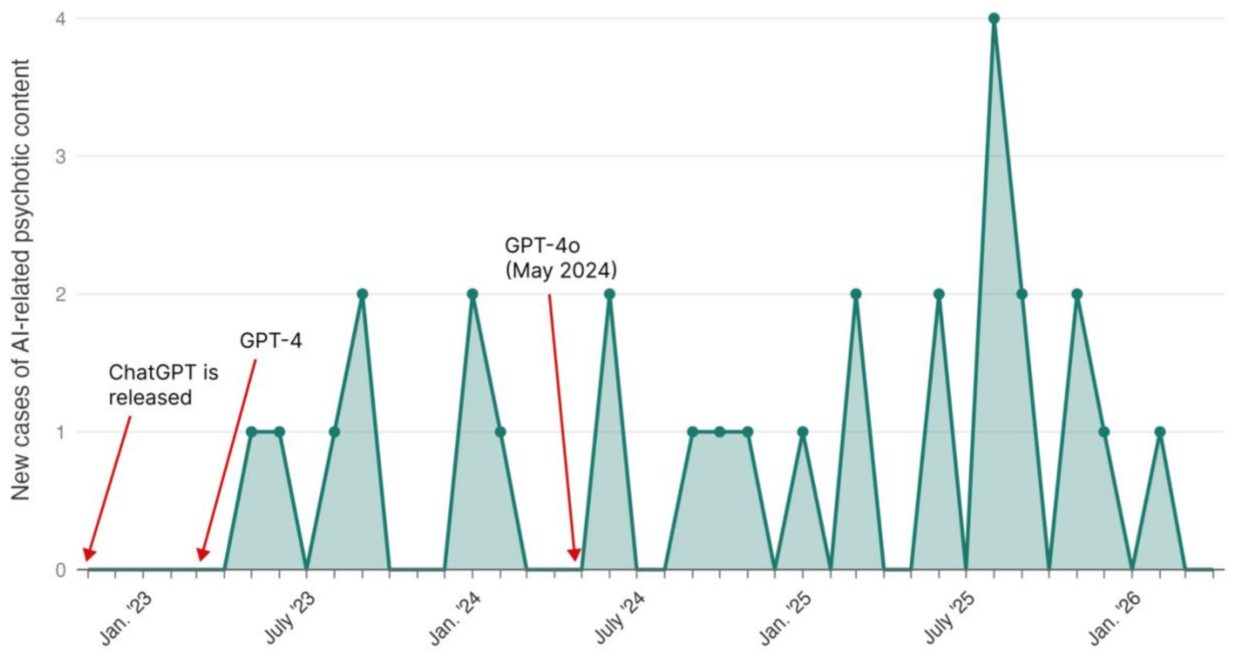

**Supplementary Figure 2.** Tracking documentation of neutral AI interactions in patients following ChatGPT's release on November 30, 2022 ($n = 17$)

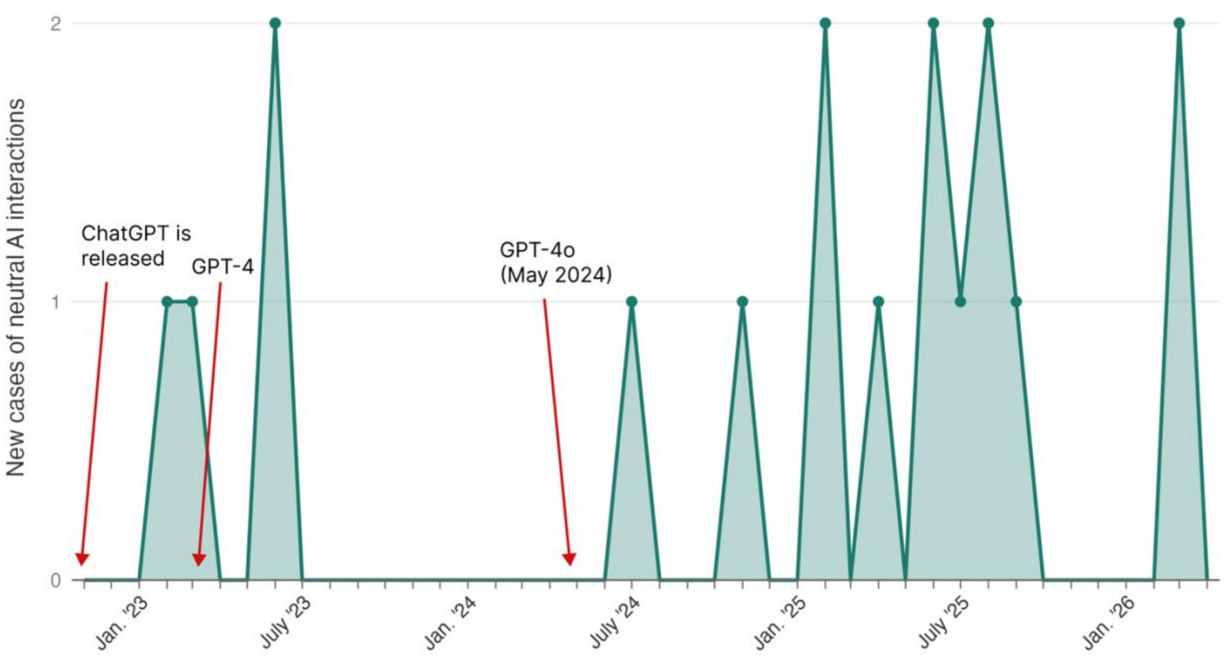
